# Neural Correlates of Positive and Negative Formal Thought Disorder in Individuals with Schizophrenia: An ENIGMA Schizophrenia Working Group Study

**DOI:** 10.1101/2023.06.06.23291034

**Authors:** Rachel J. Sharkey, Chelsea Bacon, Zeru Peterson, Kelly Rootes-Murdy, Raymond Salvador, Edith Pomarol-Clotet, Andriana Karuk, Philipp Homan, Ellen Ji, Wolfgang Omlor, Stephanie Homan, Foivos Georgiadis, Stefan Kaiser, Matthias Kirschner, Stefan Ehrlich, Udo Dannlowski, Dominik Grotegerd, Janik Goltermann, Susanne Meinert, Tilo Kircher, Frederike Stein, Katharina Brosch, Axel Krug, Igor Nenadić, Kang Sim, Gianfranco Spalletta, Fabrizio Piras, Nerisa Banaj, Scott R Sponheim, Caroline Demro, Ian S Ramsay, Margaret King, Yann Quidé, Melissa J. Green, Dana Nguyen, Adrian Preda, Vince D. Calhoun, Jessica A. Turner, Theo G.M. van Erp, Thomas Nickl-Jockschat

## Abstract

Formal thought disorder (FTD) is a key clinical factor in schizophrenia, but the neurobiological underpinnings remain unclear. In particular, relationship between FTD symptom dimensions and patterns of regional brain volume deficiencies in schizophrenia remain to be established in large cohorts. Even less is known about the cellular basis of FTD. Our study addresses these major obstacles based on a large multi-site cohort through the ENIGMA Schizophrenia Working Group (752 individuals with schizophrenia and 1256 controls), to unravel the neuroanatomy of positive, negative and total FTD in schizophrenia and their cellular bases. We used virtual histology tools to relate brain structural changes associated with FTD to cellular distributions in cortical regions. We identified distinct neural networks for positive and negative FTD. Both networks encompassed fronto-occipito-amygdalar brain regions, but negative FTD showed a relative sparing of orbitofrontal cortical thickness, while positive FTD also affected lateral temporal cortices. Virtual histology identified distinct transcriptomic fingerprints associated for both symptom dimensions. Negative FTD was linked to neuronal and astrocyte fingerprints, while positive FTD was also linked to microglial cell types. These findings relate different dimensions of FTD to distinct brain structural changes and their cellular underpinnings, improve our mechanistic understanding of these key psychotic symptoms.

## Introduction

Formal thought disorder (FTD) is a syndrome characterized by disorganized and incoherent speech [1], [2]. While FTD is a cross-diagnostic syndrome, it constitutes a key clinical factor of schizophrenia and a core component of the diagnostic criteria for this disorder in the DSM-5. Multiple lines of evidence point towards a key role for FTD in the pathophysiology of schizophrenia. It predicts transition into psychosis in clinical high-risk samples[3]–[5] and is closely correlated with long-term outcome in chronic disease states [6]–[11]. Clinically, FTD is highly heterogeneous, with impairments ranging from impoverished thought to disorganized thinking to pressured speech [12]. These different facets differentially impact clinical outcomes [7], [9], [13]. To reduce this heterogeneity, researchers have suggested grouping FTD in positive and negative symptoms [14]–[16]. While positive formal thought disorder symptoms are characterized by disorganized or unusual forms of thought or language, negative formal thought disorder includes paucity or slowing of thought or speech [12].

Brain volume reductions in fronto-temporo-basal ganglia-thalamic networks are a hallmark of schizophrenia pathology [17], [18] and might provide a major avenue towards unraveling the neurobiological basis of FTD. These brain structural abnormalities are progressive over the disease course [17] and are, at least retrospectively, linked to symptom patterns [19], [20]. Although there is an established relationship between volume deficits and overall symptom patterns in schizophrenia, major questions remain, specifically about the neuroanatomical basis of FTD in schizophrenia [18], [19], [21]. Several pioneering publications reported brain structural correlates of FTD, but these studies enrolled comparatively small numbers of schizophrenia patients (20-30 patients) [22]–[24]. Studies on much larger samples of thousands of patients, on the other hand, have pooled across patients with various diagnoses [10], [25], [26]. This approach reduces the risk of spurious correlations and increases the power compared to smaller samples; it also has the advantage that it can identify transdiagnostic markers of FTD. However, it might be less sensitive towards pathologies associated specifically with schizophrenia, as their progressive nature, their location and extent might set them apart from brain structural changes observed in other disorders [17], [27], [28].

Finally, a major limitation of conventional structural neuroimaging is that its findings can reflect various histological correlates and are, necessarily, neurobiologically nonspecific. Prior work in *post-mortem* samples has suggested that multiple neuronal and glial cell types are likely involved in the pathophysiology of schizophrenia, making histological specificity especially important [29]–[32]. The lack of specificity in structural neuroimaging is a major obstacle with regard to the identification of cellular mechanisms and neuronal circuits associated with FTD, which is difficult to model in animals. However, while neuroimaging even at ultra-high magnet strengths cannot provide a sufficient resolution to capture this level of detail, modern computational approaches based on gene expression data allow inference on the cellular composition of brain regions [33], [34]. These approaches have been subsumed under the umbrella term “virtual histology”.

This study assesses the neuroanatomical basis of FTD based on a large multi-site cohort and uses virtual histology tools on implicated brain regions to assess its cellular underpinnings. It presents the first study using a large, multi-site ENIGMA Schizophrenia Working Group dataset to identify the structural correlates of FTD. As previous neuroimaging studies [35], [36] have suggested at least at distinct correlates of positive and negative FTD, we decided to separately explore different symptom dimensions, namely positive, negative and total formal thought disorder. We then use virtual histology tools to relate each of the structural findings to cellular distributions in the cortex. To our knowledge, this is the largest study on the structural correlates of FTD in schizophrenia to date, and the first to apply virtual histology to the structural correlates FTD.

## Results

### Structural Associations with Formal Thought Disorder

We employed a general linear model to associate FTD scores with regional surface area or cortical thickness measurements of each Desikan-Killiany brain atlas region [37], controlling for age, age squared to capture potential non-linear relationships, gender and intracranial volume with FDR correction. Separate models were used for total (PANSS items: P2, N5, N6, N7), positive (P2) and negative (N5, N6, N7) FTD, and also for each of the three PANSS items contributing to negative FTD separately. These definitions of formal thought disorder have been used previously for investigating the neural basis of formal thought disorder [35].

### Total Formal Thought Disorder

Total FTD was associated with lower volumes of the bilateral pallidum and left amygdala, and thicker cortex in fronto-occipital regions. In addition, association with thicker cortex in rostral middle frontal and postcentral pole regions, as well as right caudal anterior cingulate cortex and lateral occipital and pericalcarine regions were implicated (Figure 1A, D, G; Table 1).

**Figure 1.**
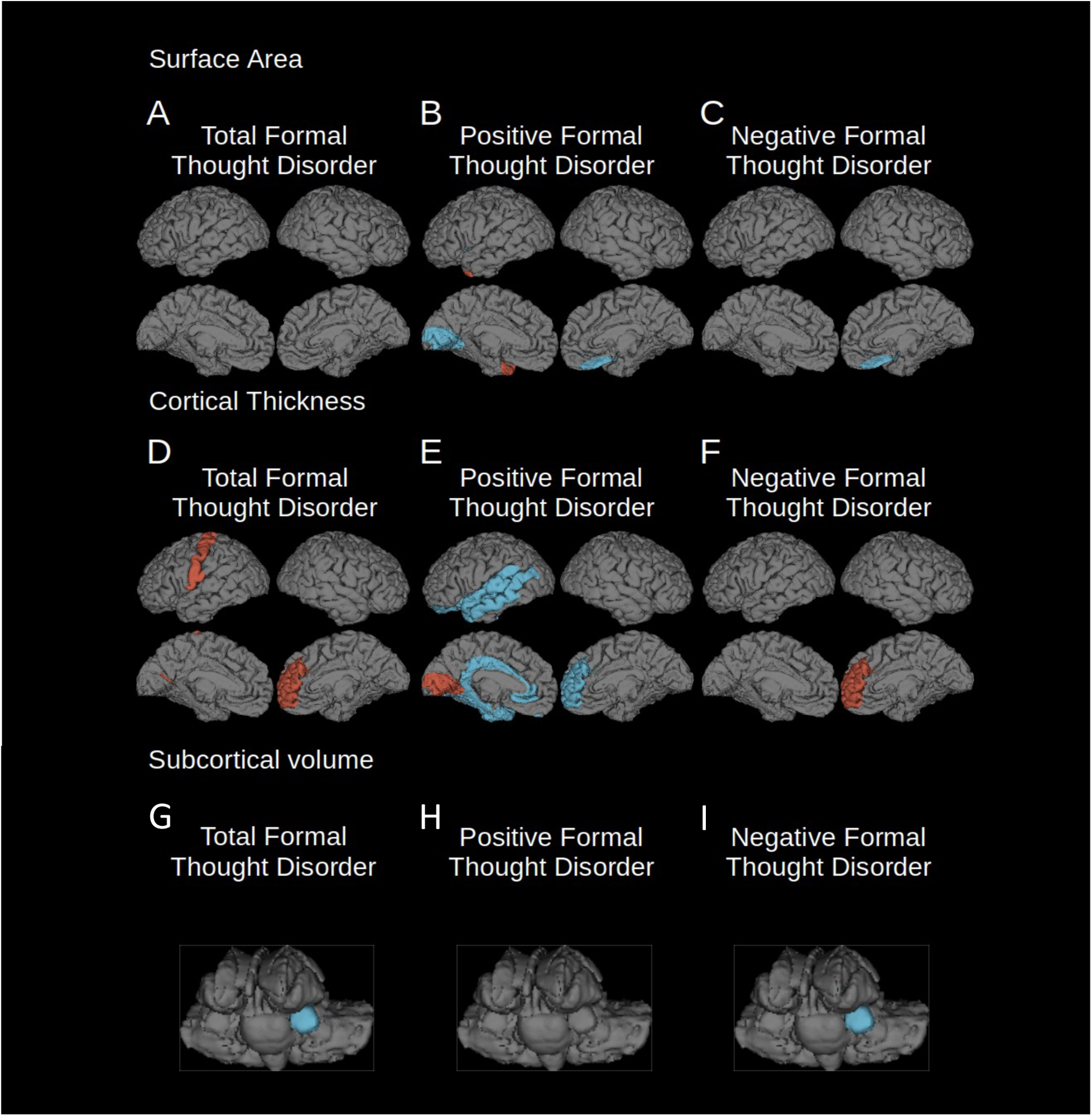
Regions where cortical surface area or thickness were significantly associated with total, positive and negative FTD (FDR corrected p = 0.001). Left hemisphere is shown on the left of each group of images. Blue indicates regions of reduced surface area, cortical thickness or subcortical volumes, red indicates regions of relatively spared surface area, cortical thickness or subcortical volumes. A) surface area changes associated with total FTD; B) surface area changes associated with positive FTD; C) surface area changes associated with negative FTD; D) cortical thickness changes associated with total FTD; E) cortical thickness changes associated with positive FTD; F) cortical thickness changes associated with negative FTD; G) volume changes of subcortical structures associated with total FTD; H) volume changes of subcortical structures associated with positive FTD; I) volume changes of subcortical structures associated with negative FTD.

**Table 1.**
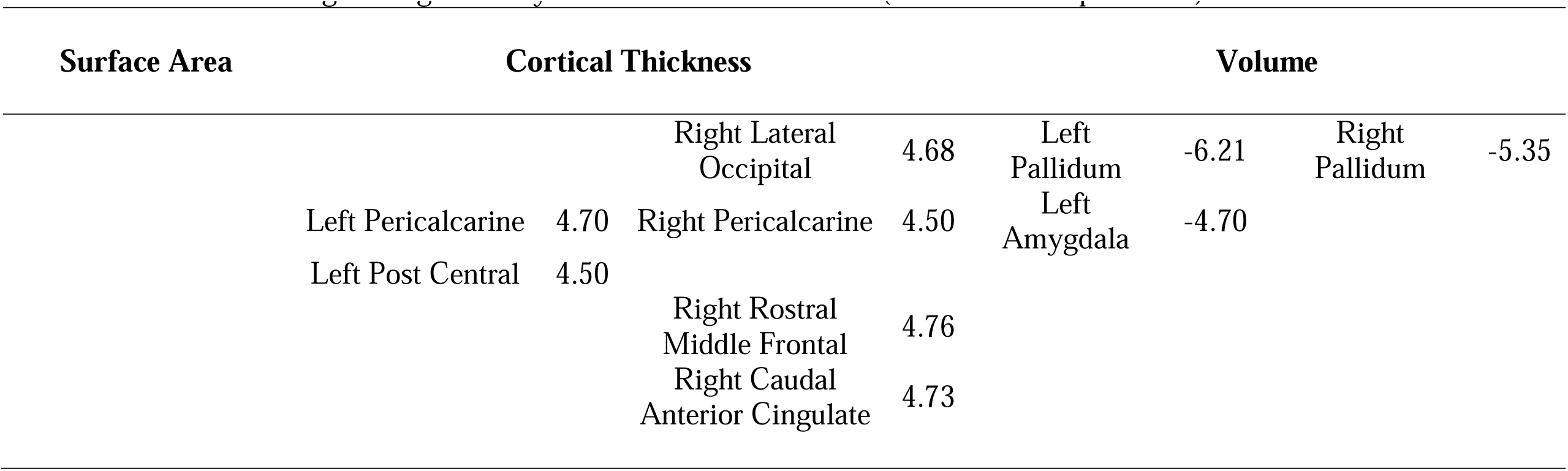
T statistics of regions significantly associated with total FTD (p = 0.001, FDR corrected).

### Positive Formal Thought Disorder

Positive FTD was associated with frontal, temporal and occipital cortical regions. These include associations with smaller surface area in the bilateral temporal, bilateral lingual, and right medial orbitofrontal cortex, and associations with thinner cortex in in the left lateral orbitofrontal and right medial orbitofrontal cortex, the rostral anterior cingulate, left caudal anterior and posterior cingulate, and temporal cortex (including the superior and middle temporal gyri and temporal poles), the parahippocampal, entorhinal and fusiform cortex, and the insula. There were also associations with thicker cortex in the right cuneus, pericalcarine and lateral occipital cortex, left lingual, and pericalcarine (Figure 1B, E, H; Table 2).

**Table 2.**
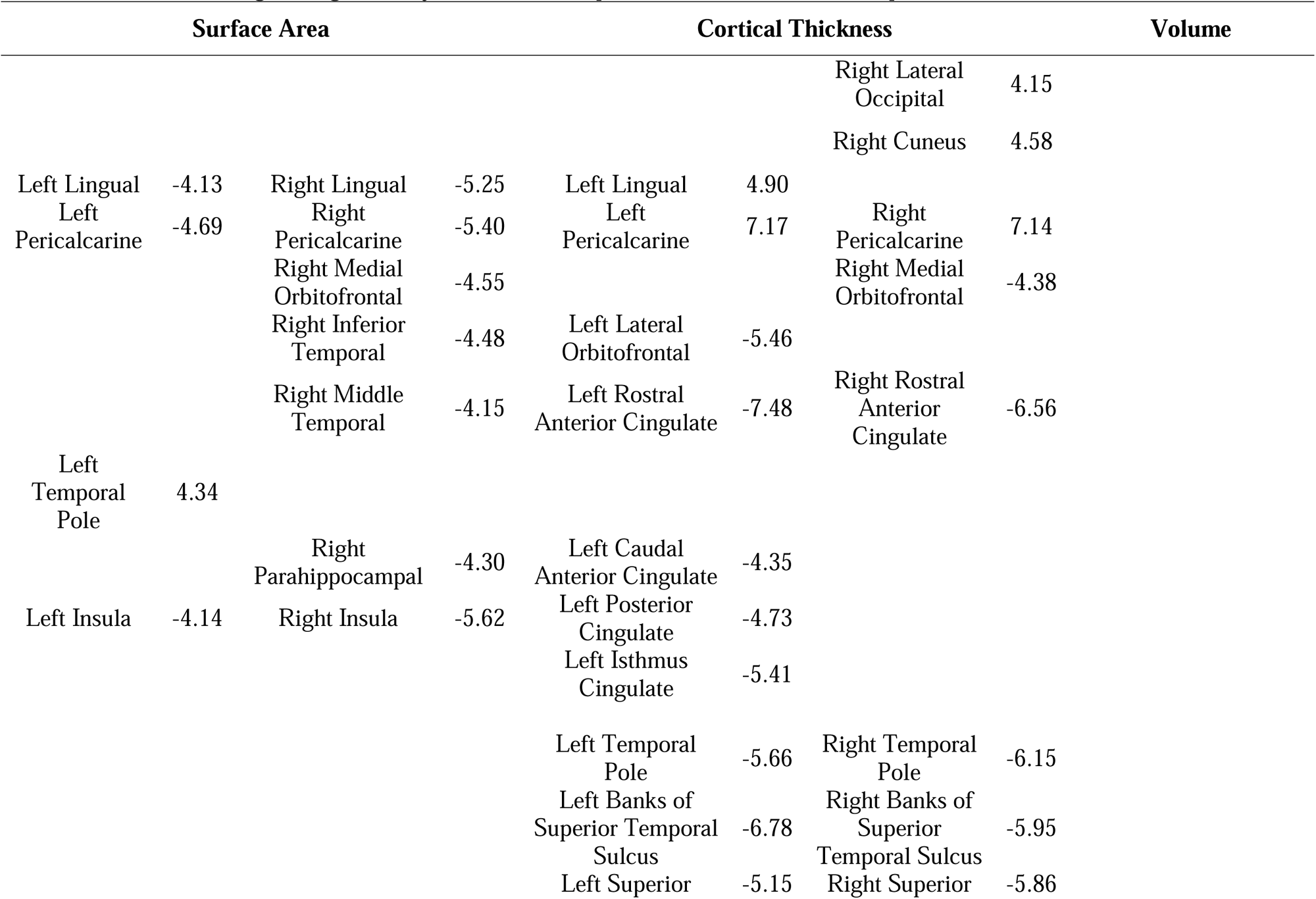
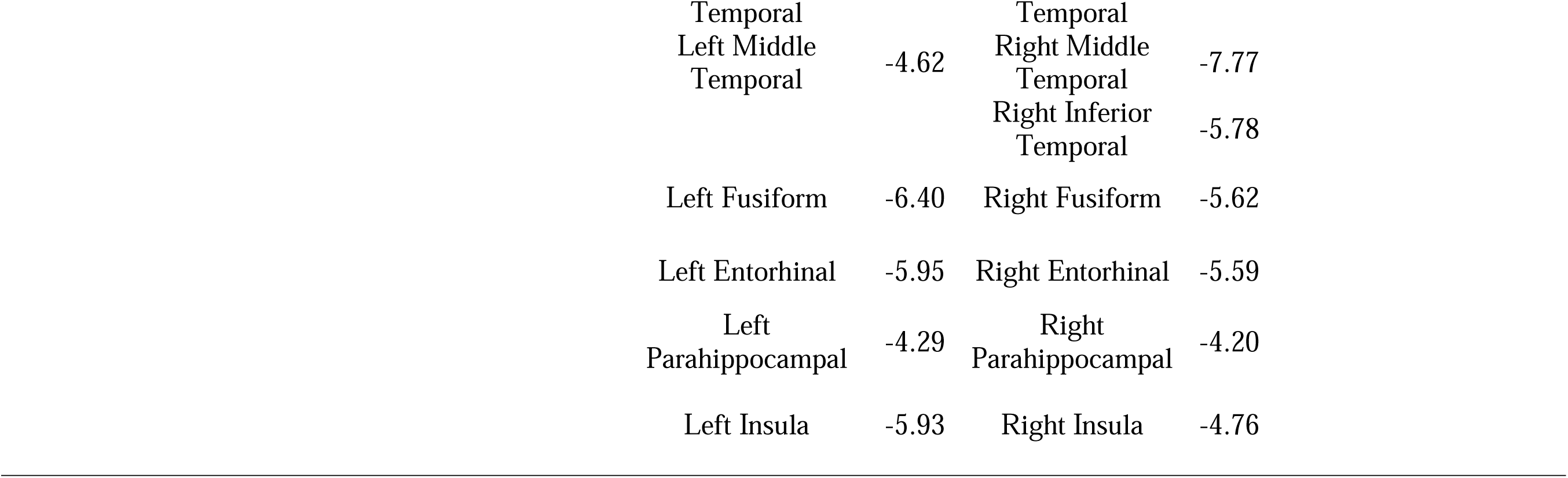
T statistics of regions significantly associated with positive FTD (p = 0.001, FDR corrected).

### Negative Formal Thought Disorder

Negative FTD was associated with smaller surface area in the lateral occipital cortex, lower pallidum and left amygdala volume, and thicker cortex in the right caudal anterior cingulate and rostral middle frontal gyri. Examining the three PANSS scores contributing to the negative FTD score separately, show that the significant findings were predominantly driven by associations with item N5 (difficulty with abstract thinking) ratings. Item N6 ratings were not significantly associated with cortical volume, and item N7 ratings were only associated with thicker medial orbitofrontal cortex (Figure 1C, F, I; Table 3; Supplementary Table 3).

**Table 3.**
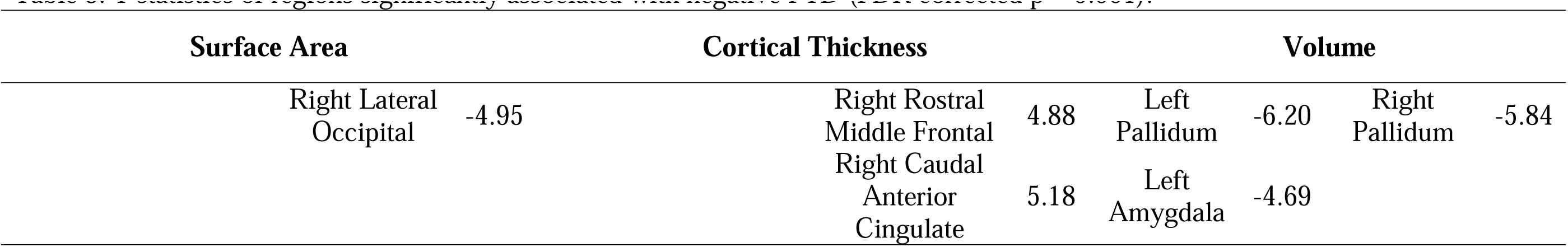
T statistics of regions significantly associated with negative FTD (p = 0.001, FDR corrected).

**Table 4.**
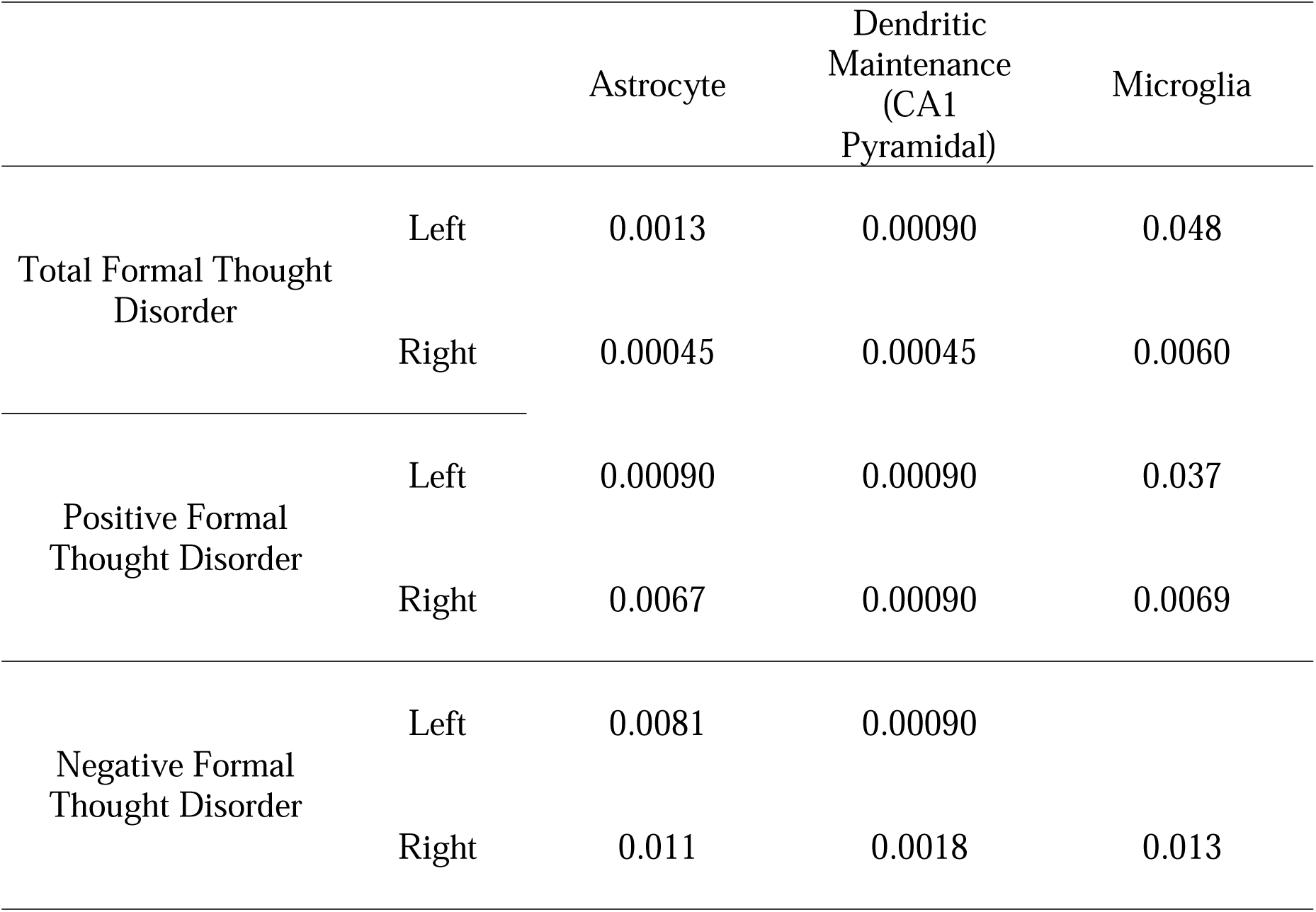
FDR Corrected P-Values for associations between FTD related cortical thickness changes and distributions of cellular transcriptomic signatures.

|  |  | Astrocyte | Dendritic Maintenance<br>(CA1 Pyramidal) | Microglia |
| --- | --- | --- | --- | --- |
| Total Formal Thought Disorder | Left | 0.0013 | 0.00090 | 0.048 |
|  | Right | 0.00045 | 0.00045 | 0.0060 |
| Positive Formal Thought Disorder | Left | 0.00090 | 0.00090 | 0.037 |
|  | Right | 0.0067 | 0.00090 | 0.0069 |
| Negative Formal Thought Disorder | Left | 0.0081 | 0.00090 |  |
|  | Right | 0.011 | 0.0018 | 0.013 |

While a fronto-occipital network was implicated in all three symptom dimensions–total, positive and negative FTD–frontal changes, in particular those in the medial orbitofrontal cortex, showed opposite patterns for cortical thickness: positive FTD was associated with *thinner* cortex, while total and negative FTD was associated with *thicker* cortex relative to other patients. Lateral temporal regions were only implicated in positive FTD. These findings suggest a neurobiological divergence of positive and negative FTD and are in line with recent functional neuroimaging findings [21], [35].

### The influence of site on the results

To assess possible site-specific effects on the results, we conducted a leave-one-out analysis. Results were consistent for positive and negative FTD, while findings for total FTD did not reproduce (Figure 2, Tables 5 and 6). These results suggest robust findings for both FTD subscales, while structural correlates of total FTD may at least partially vary by site.

**Figure 2.**
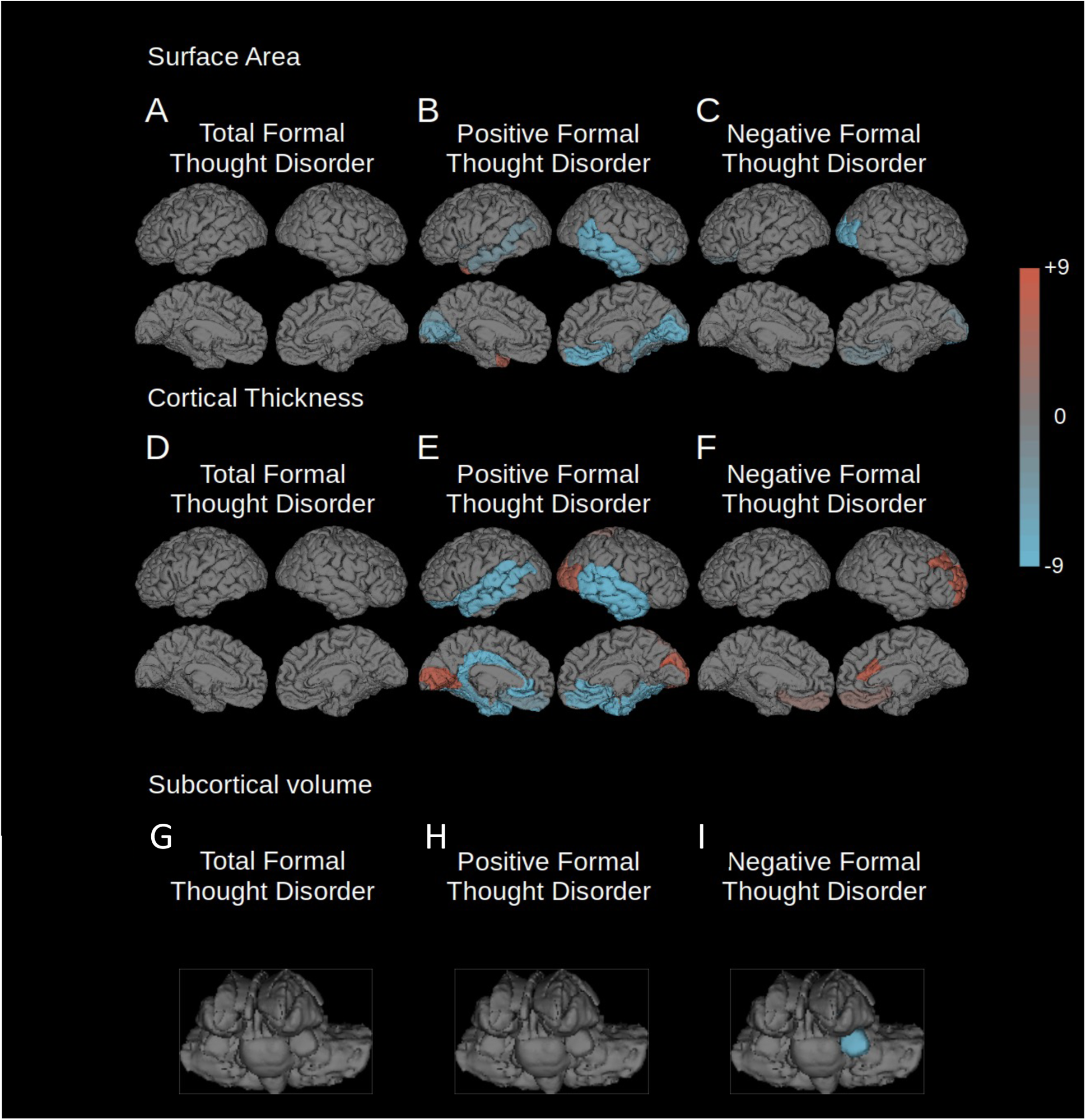
“Leave one out” analysis of FTD correlation. The regions associated with total, positive, or negative FTD (FDR corrected p = 0.001) of each of the 10 “leave one out” analyses were added together so that all positively affected regions are shown in red, and all negatively affected regions in blue. The intensity of the color represents the number of “leave one out” analyses where that region was affected. Leave one out analyses were conducted for the following parameters: A) surface area changes associated with total FTD; B) surface area changes associated with positive FTD; C) surface area changes associated with negative FTD; D) cortical thickness changes associated with total FTD; E) cortical thickness changes associated with positive FTD; F) cortical thickness changes associated with negative FTD; G) volume changes of subcortical structures associated with total FTD; H) volume changes of subcortical structures associated with positive FTD; I) volume changes of subcortical structures associated with negative FTD.

**Table 5.**
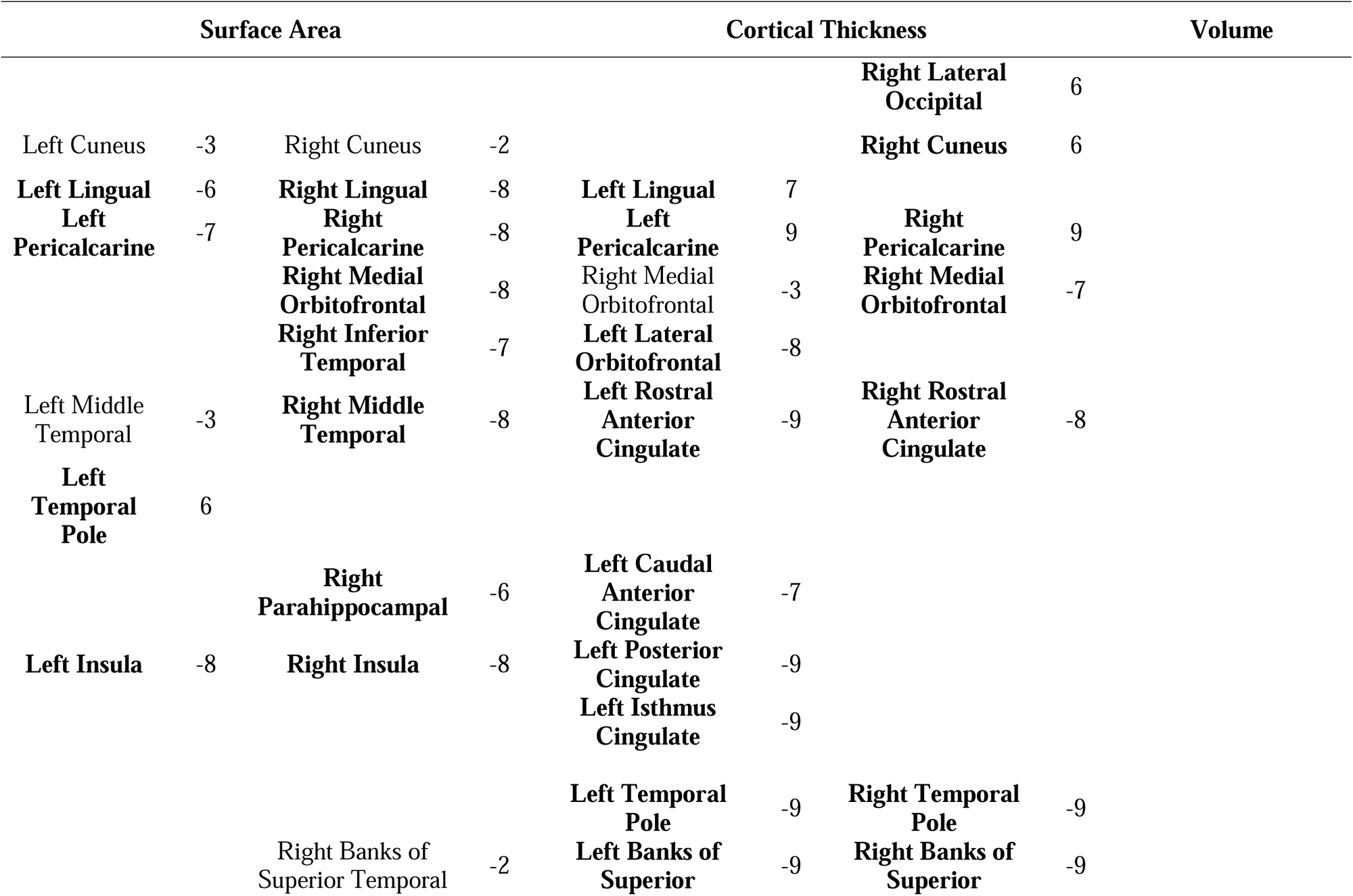
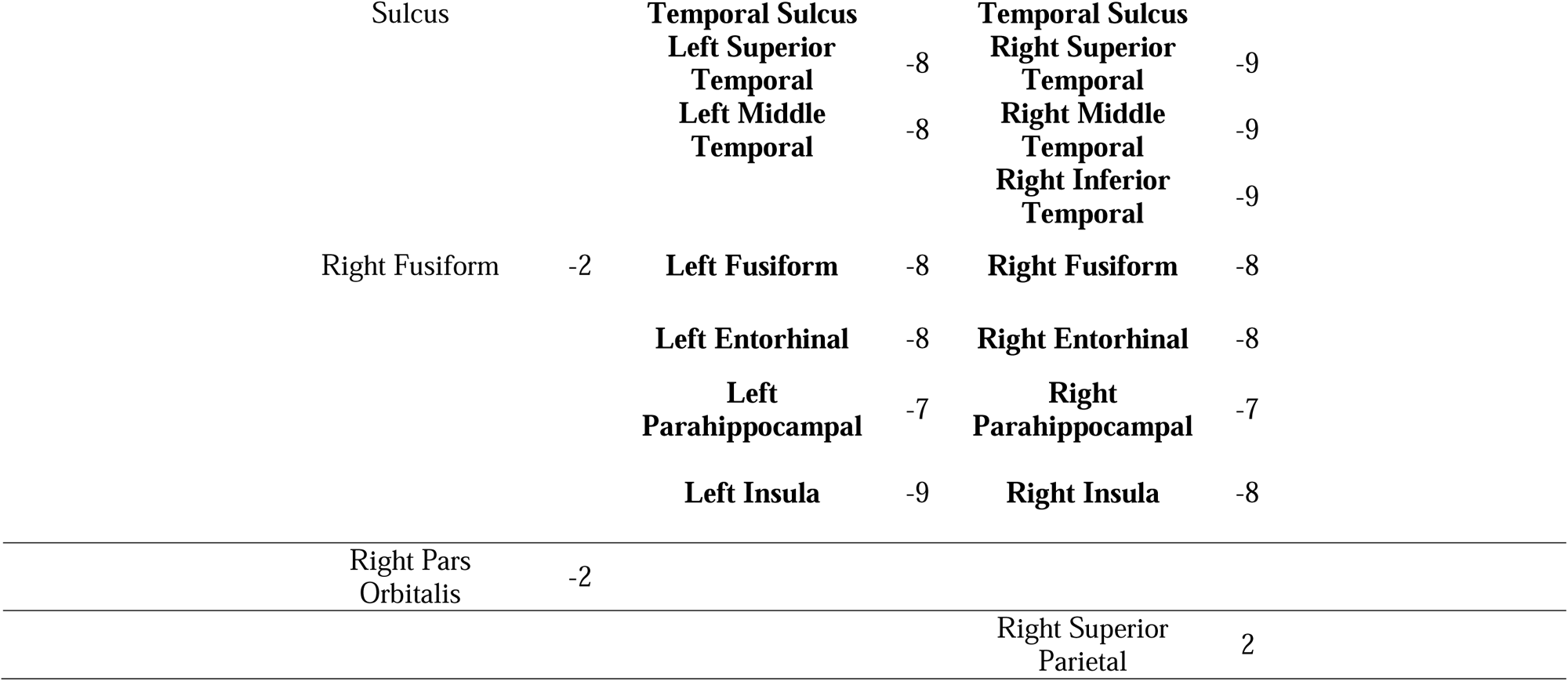
Results of the leave-one-out (LOO) analyses for positive FTD. All regions are displayed that were significant for at least one LOO analysis (p = 0.001, FDR corrected). Numbers provided indicate the number of times that these regions were retrieved as significant during the individual LOO runs. Positive numbers indicate positive correlations with FTD, negative numbers indicate negative correlations. Regions that were retrieved by the main analysis are displayed in bold.

**Table 6.**
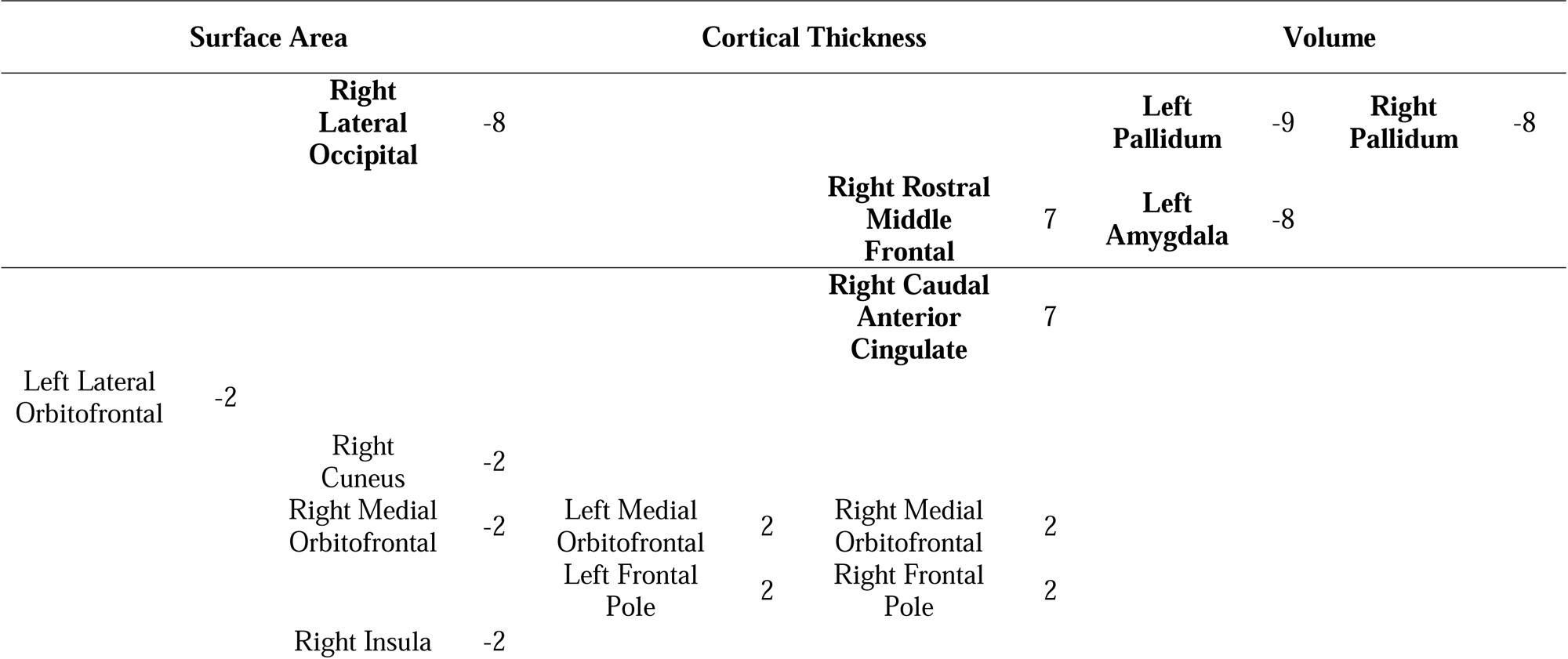
Results of the leave-one-out (LOO) analyses for negative FTD. All regions are displayed that were significant for at least one LOO analysis (p = 0.001, FDR corrected). Numbers provided indicate the number of times that these regions were retrieved as significant during the individual LOO runs. Positive numbers indicate positive correlations with FTD, negative numbers indicate negative correlations. Regions that were retrieved by the main analysis are displayed in bold.

### Patient vs. Control Comparisons

In order to compare results related to FTD with those more generally associated with schizophrenia, we used a two-sample t-test comparing patients with schizophrenia with controls. We identified wide-spread smaller cortical surface area and volume, smaller subcortical volumes, and larger lateral ventricle volumes in patients compared to healthy controls, but also larger left caudate volumes in schizophrenia. The regions that showed positive associations with FTD were smaller in schizophrenia compared to the control group, indicating a process of relative sparing rather than enlargement (Supplementary Table 2).

### Auditory Hallucination Associations

Prior research has led to the hypothesis that FTD primarily emerges from disturbances in language processing networks (dyssemantic hypothesis of FTD) [38], [39]. To test whether the identified structural changes were specific for FTD or rather indicative of unspecific changes in language processing networks in schizophrenia [2], we explored potential correlations between structural variation in FTD-related brain regions and auditory hallucinations, another major language-associated pathology in schizophrenia [40]. The regions which were significantly associated with FTD were also examined for a relationship with auditory hallucination symptoms using similar general linear models. There were no significant relationships with auditory hallucinations, suggesting the observed network is specific to FTD, not general to auditory and language dysfunction in schizophrenia.

### Cellular Genetic Fingerprint Associations

MRI imaging has provided valuable insight into *in vivo* pathologies in schizophrenia, but is unable to provide information about underlying histological changes. Novel virtual histology approaches based on gene expression databases such as the Allen Human Brain Atlas [41] utilize complex gene expression patterns to identify the cellular composition of a given brain region [33], [34]. Capitalizing on this approach, we correlated the distribution of these gene expression fingerprints with the patterns of cortical thickness change identified in our data as described previously [33]. All FTD dimensions were bilaterally associated with transcriptomic fingerprints for astrocytes (“astrocyte”) and dendritic spine maintenance (“CA1 pyramidal”). The “CA1 pyramidal” label here refers to the original source for the genetic fingerprint, but the respective gene expression patterns have been confirmed to be distributed throughout the cortex [33]. Positive FTD was also associated with the fingerprint for microglial function (“microglia”), but negative FTD was associated with microglial function in the right hemisphere only. Hence, the brain structural dissociation between positive and negative FTD appears to be somewhat accompanied by a dissociation also on a cellular level (Table 4). Positive FTD, which was associated with greater cortical atrophy was also more strongly associated with microglia which have been previously associated with excessive synaptic pruning in schizophrenia [42].

## Discussion

This study identified novel neural networks associated with different symptom dimensions of FTD in a large cohort of individuals with schizophrenia (Figure 1). While prior studies of FTD, including meta-analyses have identified abnormalities in superior temporal gyrus activation [43] and connectivity associated with FTD–the latter specifically for positive FTD [35]. This study suggests that these earlier findings are only one component of a more extensive network. The temporal lobe component of the structural network identified in this study overlaps with previously identified functional findings but connects to a wider range of regions, especially in the occipital lobe which is rarely studied in the context of schizophrenia. Both positive and negative FTD were found to be related to fronto-occipital brain regions, namely the medial orbitofrontal cortex, anterior cingulate, lateral occipital cortex and negative FTD was also found to be related to the left amygdala. However, anatomical measures related differentially between the two FTD dimensions. The associations of our FTD core network provide further insight into a long-standing controversy in the field: whether FTD emerges from dysfunction in language processing networks (“dyssemantic hypothesis”) [44] or rather from deficits in higher-order cognitive processes (“dysexecutive hypothesis”) [38]. Of note, these core regions were outside canonical language-related circuits [45], but rather associated with cognitive and behavioral control (*medial orbitofrontal* [46]–[48] and *anterior cingulate* [49], [50]), affective processing (*amygdala* [51], [52]) which have been associated with schizophrenia in previous studies [18], [22], [53], and abstract thinking and imagination (*lateral occipital cortex* [54]–[59]), which has been less commonly associated with schizophrenia [57], [58], [60]. Our findings suggest a potential role for dysfunctional executive processing as a common feature shared across FTD domains. However, it should be noted that positive FTD was also associated with classical language-related regions, suggesting a role for impaired semantic functions especially in the case of positive FTD (Figure 1B, 1E; Tables 1 – 3).

A closer look at differences between findings for distinct FTD symptom dimensions demonstrates a dissociation between the structural features of positive and negative FTD. In particular, positive and negative FTD showed opposing patterns of associations with cortical thickness in the orbitofrontal cortex and the rostral anterior cingulate (Figure 1B-C, 1E-F). Negative FTD showed a positive correlation with cortical thickness in these frontal brain regions, however, it should be noted that these findings were indicative of a relative sparing from a fronto-temporal pattern of atrophy [18], [22] in individuals with schizophrenia, rather than an absolute increase, when compared to healthy controls (Figure 1). When schizophrenia subjects were compared to healthy controls, widespread smaller cortical surface area, thinner cortex, and smaller volumes were observed (Supplementary Table 2). Different deficiency patterns in two central hubs involved in cognitive control may indicate that differential biological mechanisms or different cellular populations play a role in the emergence of these types of FTD. Additionally, positive FTD was the only symptom dimension that implicated brain regions in the temporal cortex, particularly in language-related areas (Figure 1B, 1E; Table 2). A previous meta-analysis from our lab highlighted functional changes of the superior and medial temporal gyrus in FTD [43]; central hubs of the human language processing network [61]. The temporal pole, in turn, has been linked to a semantic network involved in creative thinking [62]. Importantly, connectome-based modeling with seeds in the superior and medial temporal cortex edpredicted positive FTD symptom severity, but not any other FTD dimension [21], which is well in line with our own findings. Together, these findings paint the picture of a role for language-related networks exclusively in positive FTD. The results of our study further support the idea of a fundamental neurobiological divergence between positive and negative symptom dimensions [1], [63] which has both been shown for general schizophrenia psychopathology and its neural correlates [21].

Due to its limited spatial resolution, MR imaging does not allow a direct link between macroscopic changes and underlying molecular or cellular pathologies, a key requirement for the development of new therapeutic approaches. Novel methods, however, allow at least indirect inference on these molecular processes. Our virtual histology approach [33], based on gene expression patterns provided by the Allen Human Brain Atlas [41], identified distinct transcriptomic fingerprints associated with each of the three symptom dimensions (Table 4). Common to both the positive and negative FTD dimensions was a transcriptomic signature associated with dendritic spine maintenance and astrocytes. Consistent with this finding, post mortem studies have reported lower dendritic spine density and impaired dendritic plasticity in the brains of individuals with schizophrenia [32], [64]. Mechanistically, loss of dendritic spines has been linked to altered function in human complex 4 (C4) [65]. Complex genetic variation in the C4 gene, in turn, has been linked to schizophrenia risk [66]. Our own finding that neuroanatomical variation associated with both FTD dimensions is situated in brain regions with a high demand for dendritic spine maintenance appears plausible in light of these prior findings. Besides their role in synapse formation during development [67], astrocytes are known to modulate glutamatergic signaling [29], [31]. Pharmacological antagonization of glutamate signaling, in turn, has been shown to induce both positive and negative FTD in healthy subjects [68]–[70]. Beyond these signatures, positive FTD was also found to be associated with brain regions enriched for another non-neuronal cellular fingerprint: microglia. As resident immune cells of the central nervous system, microglia cells are involved in synaptic pruning during development [42], [71].

Our study has several limitations. The large sample size of our study made possible by the ENIGMA consortium has enabled us to identify novel neural networks associated with FTD that earlier, smaller studies were unable to identify. However, pooling over multiple sites limited us to operationalization of FTD through the use of several PANSS items, as rating scales tailored more precisely for formal thought disorder were not available across all cohorts. Additionally, neuroanatomical abnormalities in schizophrenia have been shown to be progressive. Hence, a longitudinal approach might provide an even better link between brain structural alterations and FTD than a cross-sectional approach and will be an important aim for future studies. Finally, we used a recently established virtual histology approach to identify potential cellular contributions to FTD [33]. While we regard this approach as a unique option to generate observer-independent data-driven hypotheses about the cellular underpinnings of brain structural changes associated with FTD, it does not provide direct evidence of cellular pathologies in the brains of individuals with schizophrenia. Postmortem histological studies with ante-mortem FTD ratings are warranted to verify or falsify the virtual histology findings.

In sum, this study demonstrates a convergence between neuroimaging and cellular endophenotypes and is, to the best of our knowledge, the first to associate glial function with formal thought disorder specifically. The identification of a multi-scale associations between structural and transcriptomic networks associated with cellular function is of specific interest clinically, because it provides the basis for linking neuroimaging findings and clinically relevant molecular targets in a way that is not possible with either method in isolation.

## Methods

### ENIGMA Schizophrenia Working Group Data

This study included the data from 752 individuals with schizophrenia and 1256 healthy controls pooled by the ENIGMA Consortium Schizophrenia Working Group. The data comprised cortical thickness and cortical surface area, and subcortical volume for each region in the Desikan-Killiany atlas, subcortical volumes, demographic information, and PANSS symptom scores (see Supplementary Table 1 for sample details and image processing).

### Formal Thought Disorder Scores

We calculated formal thought disorder and auditory hallucination symptom score for each patient from the PANSS. Four symptom scores were calculated for each patient: total formal thought disorder, positive formal thought disorder, negative formal thought disorder and hallucinations. The total formal thought disorder scale was derived from the sum of items P2, N5, N6 and N7, as has been used in previous studies [35]. Of this subset, P2 was used as a measure of positive formal thought disorder, while negative formal thought disorder was measured as the sum of N5, N6 and N7. Hallucinations were measured by the P3 score. Tests of the relationship between formal thought disorder related findings and auditory hallucinations were carried out to test the specificity of the results to formal thought disorder.

### General Linear Models

Each of the major comparisons in this study used a general linear model examining associations between a variable of interest and the regional cortical thickness, surface area and subcortical volume values in a mass univariate manner which was then controlled for multiple comparisons to false discovery rate q=0.001. Each model controlled for age, age squared, gender and intracranial volume in format:

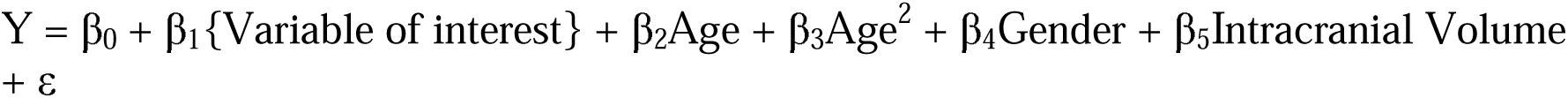

This model was used for the relationship between cortical thickness and surface area and subcortical volume and: Total Formal Thought Disorder, Positive Formal Thought Disorder, Negative Formal Thought Disorder, N5, N6, N7 and Diagnosis. The same model was applied to the relationship between Auditory Hallucinations for regions that showed significant correlations with the formal thought disorder scores. Similar statistical models have been used in multiple ENIGMA studies [72], [73].

### Leave-one-out Validation for Site Variability

Matlab scripts were written to check for findings predominantly driven by results from single sites. This analysis repeated the statistical analysis 10 times, each time removing the data from one site. Using the output of these analyses, we summarized any regions that appeared as significant in one or more of the leave one out analyses, taking note of the directionality of the change (see Figure 3).

### Schizophrenia vs. Control Comparison

A bidirectional two-sample t-test was performed comparing the schizophrenia and control groups, without controlling for the additional variables of no interest. This was included along with the GLM for diagnosis effects to model effects which include the global reduction in ICV which has been previously demonstrated in schizophrenia.

### Cellular Genetic Fingerprint Associations

R scripts released by the Paus Lab were used to associate our regional cortical thickness findings with the regional distributions of different cellular fingerprints (see Shin et al 2018 for details on the method) [33].

## Data Availability

All data produced in the present work is available via the ENIGMA consortium.

## Acknowledgements

FIDMAG: Supported by Instituto de Salud Carlos III (Co-funded by European Regional Development Fund/European Social Fund) “Investing in your future”): Miguel Servet Research Contract (CPII16/00018 to E. Pomarol-Clotet and CP14/00041 to J. Radua.).

SWIFT (Homan, Zurich): Supported by a NARSAD grant from the Brain & Behavior Research Foundation (28445) and by a Research Grant from the Novartis Foundation (20A058).

FOR2107 Marburg: This work was funded by the German Research Foundation (DFG), Tilo Kircher (speaker FOR2107; DFG grant numbers KI 588/14-1, KI 588/14-2), Axel Krug (KR 3822/5-1, KR 3822/7-2), Igor Nenadić (NE 2254/1-2), Carsten Konrad (KO 4291/3-1).

FOR2107 Münster: This work is part of the German multicenter consortium “Neurobiology of Affective Disorders. A translational perspective on brain structure and function“, funded by the German Research Foundation (Deutsche Forschungsgemeinschaft DFG; Forschungsgruppe/Research Unit FOR2107). Work Package WP1, FOR2107/MACS cohort and brainimaging: Udo Dannlowski (co-speaker FOR2107; DA 1151/5-1, DA 1151/5-2)

IGP: The Imaging Genetics in Psychosis (IGP) study was funded by Project Grants from the Australian National Health and Medical Research Council (NHMRC; APP630471 and APP1081603), and the Macquarie University’s ARC Centre of Excellence in Cognition and its Disorders (CE110001021). This project used participants from the Australian Schizophrenia Research Bank (ASRB), funded by the NHMRC Enabling Grant (386500 held by V. Carr, U. Schall, R. Scott, A. Jablensky, B. Mowry, P. Michie, S. Catts, F. Henskens and C.Pantelis; Chief Investigators), and the Pratt Foundation, Ramsay Health Care, the Viertel Charitable Foundation, as well the Schizophrenia Research Institute, using an infrastructure grant from the NSW Ministry of Health.

RomeSL: This study was supported by grants (RC10-11-12-13-14-15/A) from the Italian Ministry of Health and by the ERANET NEURON from the European Commission.

COBRE: The COBRE project was supported by NIH grants R01EB006841 and P20GM103472, and NSF grant 1539067. Jessica A. Turner and Vince D. Calhoun are supported by 5R01MH094524 and 5R01MH121246.

Singapore: This study was supported by research grants from the National Healthcare Group, Singapore (SIG/05004), and the Singapore Bioimaging Consortium (RP C009/2006) research grants awarded to K.S.

UCISZ: The UCISZ study was supported by the National Institutes of Mental Health grant number R21MH097196 to TGMvE. UCISZ data was processed by the UCI High Performance Computing cluster supported by Joseph Farran, Harry Mangalam, and Adam Brenner and the National Center for Research Resources and the National Center for Advancing Translational Sciences, National Institutes of Health, through Grant UL1 TR000153.

T.N.-J. was supported by the Andrew H. Woods Professorship.

---

**Supplementary Table 1.** Demographics of contributions from each ENIGMA site.

**Supplementary Table 2.** Differences between regions of significantly reduced surface area, cortical thickness or subcortical volume in individuals with schizophrenia compared to controls (FTD corrected p = 0.001).

**Supplementary Table 3.** T Statistics of regions significantly associated with PANSS N5 (FDR corrected p = 0.001).

