## Supplementary Table 1 for "Neural Correlates of Positive and Negative Formal Thought Disorder in Individuals with Schizophrenia: An ENIGMA Schizophrenia Working Group Study"

| ENIGMA Sample | Number of Subjects (Number of Patients) | Age | Number of Males | Average Duration of Illness | Average PANSS Total Score (StDev) | Average PANSS Positive Score (StDev) | Average PANSS Negative Score (StDev) |
| --- | --- | --- | --- | --- | --- | --- | --- |
| FIDMAG | 279 (156) | 38.85 (11.07) | 176 | 15.97 (11.23) | 75.84 (18.39) | 16.78 (5.74) | 22.67 (6.71) |
| SWIFT | 37 (24) | 32.49 (9.32) | 22 | 9.51 (7.84) | 57.96 (12.05) | 16.38 (5.36) | 12.75 (4.99) |
| FOR2107* | 681 (40) | 32.92 (12.28) | 266 | 169.24 (119.82) | NA | NA | NA |
| IMH Singapore | 214 (138) | 32.95 (9.38) | 145 | 6.80 (7.47) | 40.16 (8.45) | 10.70 (3.79) | 9.03 (3.12) |
| RomeSL | 280 (164) | 38.60 (11.43) | 183 | 14.91 (10.69) | NA | NA | NA |
| IGP | 139 (71) | 36.35 (11.61) | 64 | 14.60 (10.16) | 45.85 (12.29) | 10.88 (6.43) | 10.32 (4.18) |
| PHCP | 128 (46) | 43.88 (13.56) | 61 | 18.55 (11.13) | NA | NA | NA |
| PENS | 51 (17) | 47.33 (9.39) | 27 | 24.71 (10.49) | NA | NA | NA |
| COBRE | 135 (65) | 36.36 (12.59) | 103 | 15.58 (12.59) | 58.71 (14.70) | 14.92 (5.24) | 14.60 (4.69) |
| UCISZ | 57 (27) | 42.11 (11.44) | 45 | 17.50 (10.05) | 59.96 (12.00) | 15.56 (4.15) | 16.04 (5.88) |

Supplementary Table 1. Demographics of contributions from each ENIGMA site. * The sample FOR2107 was collected across two sites, one at the University of Marburg, one at the University of Münster
