## Supplementary Table 2 for "Neural Correlates of Positive and Negative Formal Thought Disorder in Individuals with Schizophrenia: An ENIGMA Schizophrenia Working Group Study"

Supplementary Table 2. Differences between regions of significantly reduced surface area, cortical thickness or subcortical volume in patients with schizophrenia compared to controls (FTD corrected p = 0.001).

| **Surface Area** | | | | **Cortical Thickness** | | | | **Subcortical Volume** | | | |
| --- | --- | --- | --- | --- | --- | --- | --- | --- | --- | --- | --- |
|  |  |  |  | Left Lateral Occipital | -5.82 | Right Lateral Occipital | -5.92 | Left Lateral Ventricle | 7.76 | Right Lateral Ventricle | 6.52 |
|  |  |  |  | Left Cuneus | -5.40 | Right Cuneus | -5.90 | Left Thalamus | -5.91 | Right Thalamus | -6.27 |
|  |  |  |  | Left Lingual | -5.84 | Right Lingual | -6.57 | Left Caudate | 4.44 |  |  |
|  |  |  |  | Left Pericalcarine | -3.95 | Right Pericalcarine | -3.96 | Left Hippocampus | -5.21 | Right Hippocampus | -5.45 |
|  |  |  |  | Left Caudal Middle Frontal | -8.25 | Right Caudal Middle Frontal | -8.94 |  |  |  |  |
|  |  |  |  | Left Lateral Orbitofrontal | -4.85 | Right Lateral Orbitofrontal | -3.99 |  |  |  |  |
| Left Pars Opercularis | -4.01 | Right Pars Opercularis | -4.47 | Left Pars Opercularis | -9.15 | Right Pars Opercularis | -9.00 |  |  |  |  |
|  |  | Right Pars Orbitalis | -4.02 | Left Pars Orbitalis | -6.13 | Right Pars Orbitalis | -5.27 |  |  |  |  |
|  |  |  |  | Left Pars Triangularis | -7.06 | Right Pars Triangularis | -7.24 |  |  |  |  |
|  |  |  |  | Left Precentral | -10.41 | Right Precentral | -10.07 |  |  |  |  |
|  |  |  |  | Left Paracentral | -8.16 | Right Paracentral | -8.92 |  |  |  |  |
|  |  |  |  | Left Postcentral | -9.17 | Right Postcentral | -8.75 |  |  |  |  |
| Left Rostral Middle Frontal | -4.00 | Right Rostral Middle Frontal | -4.63 | Left Rostral Middle Frontal | -6.50 | Right Rostral Middle Frontal | -6.04 |  |  |  |  |
|  |  |  |  | Left Superior Frontal | -9.23 | Right Superior Frontal | -9.01 |  |  |  |  |
| Left Caudal Anterior Cingulate | -4.09 |  |  |  |  |  |  |  |  |  |  |
|  |  |  |  | Left Isthmus Cingulate | -4.10 | Right Isthmus Cingulate | -5.63 |  |  |  |  |
|  |  |  |  | Left Posterior Cingulate | -5.40 | Right Posterior Cingulate | -5.71 |  |  |  |  |
|  |  |  |  | Left Temporal Pole | -4.53 | Right Temporal Pole | -3.96 |  |  |  |  |
|  |  |  |  | Left Banks of Superior Temporal Sulcus | -6.68 | Right Banks of Superior Temporal Sulcus | -7.71 |  |  |  |  |
|  |  |  |  | Left Superior Temporal | -7.73 | Right Superior Temporal | -7.90 |  |  |  |  |
|  |  |  |  | Left Middle Temporal | -6.60 | Right Middle Temporal | -6.23 |  |  |  |  |
|  |  |  |  | Left Inferior Temporal | -5.56 | Right Inferior Temporal | -5.23 |  |  |  |  |
|  |  |  |  | Left Transverse Temporal | -7.23 | Right Transverse Temporal | -8.10 |  |  |  |  |
|  |  | Right Fusiform | -4.26 | Left Fusiform | -7.14 | Right Fusiform | -8.48 |  |  |  |  |
|  |  |  |  | Left Para-hippocampal | -5.56 | Right Para-hippocampal | -5.64 |  |  |  |  |
|  |  |  |  | Left Inferior Parietal | -9.01 | Right Inferior Parietal | -8.49 |  |  |  |  |
|  |  |  |  | Left Precuneus | -8.25 | Right Precuneus | -8.55 |  |  |  |  |
|  |  |  |  | Left Superior Parietal | -8.00 | Right Superior Parietal | -7.88 |  |  |  |  |
|  |  |  |  | Left Supramarginal | -9.52 | Right Supramarginal | -10.68 |  |  |  |  |
|  |  |  |  | Left Insula | -6.88 | Right Insula | -4.94 |  |  |  |  |
|  |  | Overall Right Surface Area | -4.30 | Overall Left Cortical Thickness | -10.08 | Overall Right Cortical Thickness | -10.07 |  |  |  |  |
