## Supplementary Table 3 for "Neural Correlates of Positive and Negative Formal Thought Disorder in Individuals with Schizophrenia: An ENIGMA Schizophrenia Working Group Study"

Supplementary Table 3. T Statistics of regions significantly associated with PANSS N5 (FDR corrected p = 0.001).

| **Surface Area** | | | | **Cortical Thickness** | | | | **Subcortical Volume** | | | |
| --- | --- | --- | --- | --- | --- | --- | --- | --- | --- | --- | --- |
| Left Lateral Occipital | -4.52 | Right Lateral Occipital | -5.31 | Left Rostral Middle Frontal | 4.64 | Right Rostral Middle Frontal |  | Left Pallidum | -5.77 | Right Pallidum | -6.02 |
|  |  | Right Cuneus | -5.09 |  |  |  |  | Left Amygdala | -4.55 |  |  |
| Left Lateral Orbitofrontal | -4.63 |  |  |  |  |  |  |  |  |  |  |
| Left Medial Orbitofrontal | -5.12 | Right Medial Orbitofrontal | -5.68 |  |  |  |  |  |  |  |  |
|  |  | Overall Right Surface Area | -4.66 |  |  |  |  |  |  |  |  |
